# Sex differences in profile and in-hospital mortality for acute stroke in Chile: data from a nationwide hospital registry

**DOI:** 10.1101/2023.11.22.23298935

**Authors:** Marilaura Nuñez, Ma.Ignacia Allende, Francisca Gonzalez, Gabriel Cavada, Craig S. Anderson, Paula Muñoz-Venturelli

## Abstract

**Background:** Knowledge of local contextual sex-differences in the profile and outcome for stroke can improve service delivery. We aimed to determine sex-differences in the profile of patients with acute stroke and their associations with in-hospital mortality (I-HM) in the national hospital database of Chile.

**Methods:** We present a retrospective cohort based on the analysis of the 2019 Chilean database of Diagnosis-Related Groups (DRGs), which represents 70% of the operational expenditure of the public health system. Multiple logistic regression models were used to determine independent associations of acute stroke (defined by main diagnosis ICD-10 codes) and I-HM, and reported with odds ratios (OR) and 95% confidence intervals (CI).

**Results:** Of 1,048,575 hospital discharges, 15,535 were for patients with acute stroke (7,074 [45.5%] in women) and 2,438 (15.6%) of them died in-hospital. Disparities by sex in sociodemographic and clinical characteristics were identified for stroke and main subtypes. Women with ischemic stroke had lower I-HM (OR 0.80, 95%CI 0.70-0.92; *P*=0.002) compared to men; other independent predictors included age (1.03, 1.03-1.04; *P*<0.001), chronic kidney disease (CKD) (1.48, 1.20-1.81; *P*<0.001), atrial fibrillation (1.56, 1.34-1.83; *P*<0.001), admission to hospital without a stroke unit (1.21, 1.05-1.39; *P*=0.003), and several risk factors. Conversely for intracerebral hemorrhage, women had higher I-HM than men (1.21, 1.04-1.41; P=0.02); other independent predictors included age (1.01, 1.00-1.01; *P*<0.001), CKD (1.54, 1.22-1.94; *P*<0.001), oral anticoagulant use (2.01, 1.56-2.59; *P*<0.001), and several risk factors.

**Conclusion:** Sex disparities in patients characteristics and in-hospital mortality exist for acute stroke in Chile. I-HM is higher for acute ischemic stroke in men and higher for ICH in women. Future research is need to better identify contributing factors.

## Introduction

Despite advances in knowledge and interventions, stroke remains the second leading cause of death worldwide.^1^ Further research is needed to better define determinants and risk factors to improve public health policies and a focus towards personalized medicine. Awareness is growing of sex differences in stroke, with studies consistently showing women are older at presentation, have more atrial fibrillation (AF) and hypertension (HT), and poorer functional outcomes.^2–4^ There are divergent results in regard sex disparities in case fatality,^5–8^ where most studies have been undertaken in developed countries, leaving a knowledge gap in relation to low- and middle-income countries where the influence of specific socioeconomic factors is likely to be important. Thus, our aim was to determine sex-differences in characteristics and determinants of in-hospital mortality for patients with acute stroke, using the nationwide hospital database of Chile.

## Methods

### Design

A retrospective cohort study undertaken according to STROBE criteria (see Table S1)^9^ using the Chilean International Refined Diagnosis Related Groups (DRGs) database of hospital discharges for the public health insurance scheme of Chile.^10^ DRG classification is an international standardized method of data collection from clinical discharges that categorizes patients into homogenous groups based upon similar characteristics and according to disease complexity, hospital workload, quality of service, and associated costs.^11,12^

Our analysis pertains to hospital discharges from 65 hospitals nationwide with high and medium complexity level during 2019, which represent 70% of the operating expenditure of the public health system in Chile.^12,13^ All patients (age ≥18 years) with a main diagnosis of stroke who were discharged from hospital between January 1 and December 31, 2019, and registered in the Chilean IR-DRG with complete data, were considered. In cases with multiple discharges, the second and subsequent events were included as re-admission events, except for admissions that occurred within 24 hours of last discharge which were considered as a continuous hospitalization of the first event. Patients without information on age or sex were excluded.

### Measures

Stroke diagnosis was defined as any of the following *International classification of diseases 10th version* (ICD-10) codes in the main patient diagnosis: subarachnoid hemorrhage (SAH) (I60), intracerebral hemorrhage (ICH) (I61-I62), ischemic stroke (IS) (I63) and undetermined stroke (I64).^14^ Low socioeconomic status (SES) was obtained through the national health insurance assigned group, which categorizes individuals based on their income into four main types; low SES is assigned to those without income or with an income lower than the minimum country wage.^15^ Risk factors and comorbidities were identified in secondary diagnoses by ICD-10 and in-hospital procedures by ICD-9 codes. Most variables were analyzed at the patient level, except for the presence of a stroke unit which was analyzed at the hospital level.

The main outcomes were the sex differences in patient characteristics, care and in-hospital mortality, adjusted for age, SES, risk factors, stroke subtype, and clinical characteristics. In-hospital mortality was defined as death during hospitalization that was due to stroke as the main diagnosis, in all patients discharged in 2019.

### Analysis

For bivariate analysis, categorical variables were analyzed using chi-squared independence test or Fisher’s tests. Quantitative variables were analyzed using the Student’s t test for normal distribution and Mann-Withney U test for a non-normal distribution. Sequential multivariate logistic regression models were developed with adjustments for age and then for variables of clinical relevance, such as SES and presence of a unit stroke, and the results reported as odds ratios (OR) and 95% confidence intervals (CI). A significance level was defined as *P*<0.05. All statistical analysis were performed using STATA version 17.0.

## Results

Of 1,048,575 DRGs hospital discharges in Chile during 2019, there were 15,535 patients with main diagnosis of stroke. Of these, 366 had an undetermined primary diagnosis of stroke. (Table 1, Figure 1).

**Figure 1.**
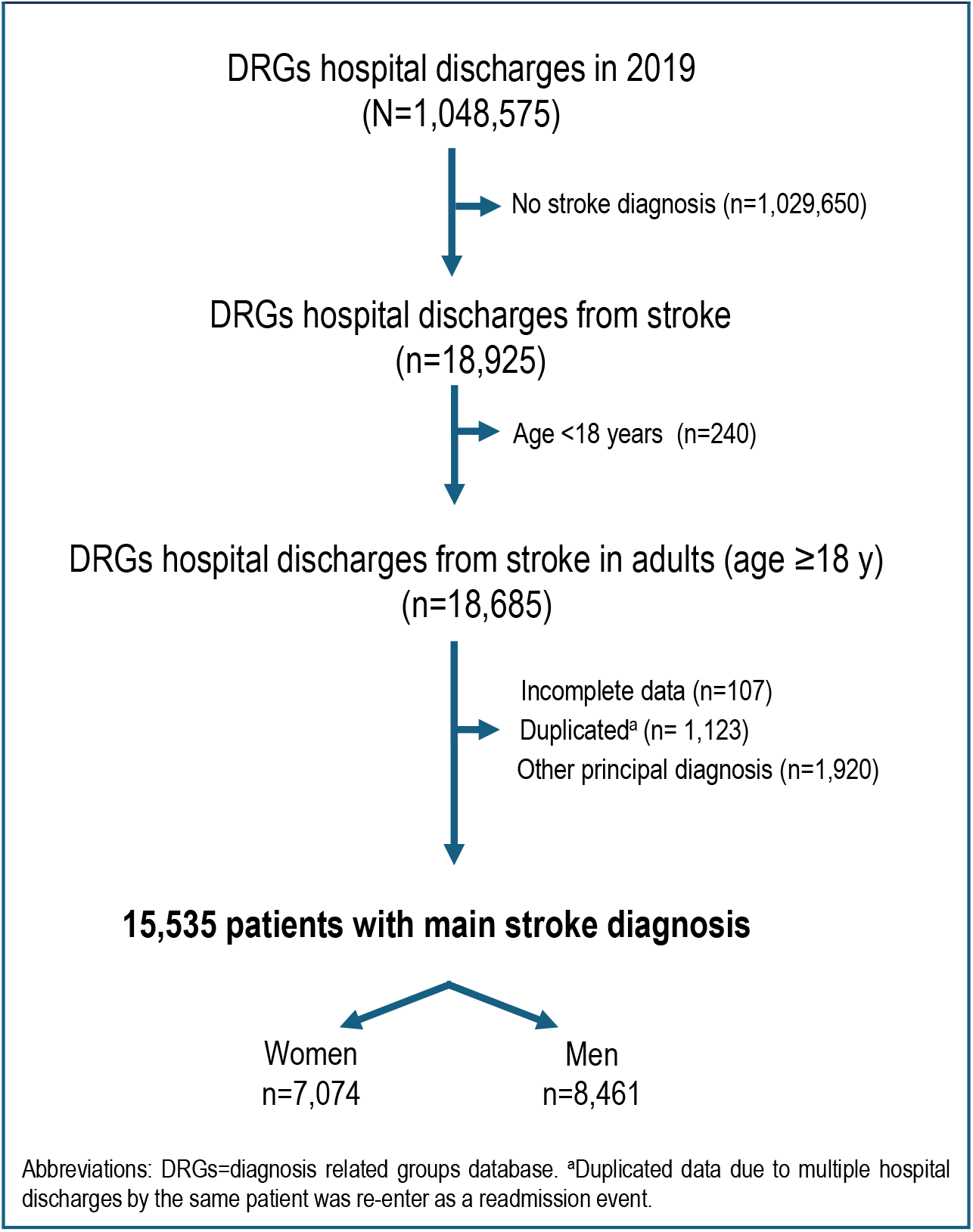
Study set-up flowchart.

**Table 1.** Baseline characteristics of overall acute stroke patients by sex and multivariable analysis of characteristics associated to women compared to men.

|  | Women<br>n=7,074; 45.54% | Men<br>n= 8,461;<br>54.46% | P Value |
| --- | --- | --- | --- |
| **Baseline characteristics, n(%)** |  |  |  |
| **Sociodemographic characteristics** |  |  |  |
| Age years, mean ± SD | 70.36 ± 14.76 | 67.00 ± 13.45 | **<0.001** |
| Socioeconomic status<sup>a</sup> |  |  | **<0.001** |
| Low | 5,783 (81.75) | 5,801 (68.56) |  |
| Middle | 565 (7.99) | 936 (11.06) |  |
| High | 572 (8.09) | 1,404 (16.59) |  |
| **Risk factors** |  |  |  |
| Hypertension | 5,477 (77.42) | 6,241 (73.76) | **<0.001** |
| Diabetes mellitus | 2,506 (35.43) | 2,860 (33.80) | **0.034** |
| Dyslipidemia | 863 (12.20) | 995 (11.76) | 0.400 |
| Current smoking | 653 (9.23) | 1,248 (14.75) | **<0.001** |
| Chronic kidney disease | 615 (8.69) | 749 (8.85) | 0.728 |
| Obesity | 617 (8.72) | 512 (6.05) | **<0.001** |
| Heart diseases | 1,818 (25.70) | 2,051 (24.24) | **0.036** |
| Ischemic heart diseases | 347 (4.91) | 711 (8.40) | **<0.001** |
| Valvular heart disease | 228 (3.22) | 201 (2.38) | **0.001** |
| Atrial fibrillation | 1,153 (16.30) | 1,086 (12.84) | **<0.001** |
| Others | 648 (9.16) | 650 (7.68) | **0.001** |
| OACs | 534 (7.55) | 509 (6.02) | **<0.001** |
| Multimorbidity<sup>b</sup> | 5,021 (70.98) | 5,805 (68.61) | **0.001** |
| **Stroke by subtype** |  |  |  |
| Ischemic stroke | 4,659 (65.86) | 5,781 (68.33) | **0.001** |
| Intracerebral hemorrhage | 1,568 (22.17) | 2,112 (24.96) | **<0.001** |
| Subarachnoid hemorrhage | 675 (9.54) | 374 (4.42) | **<0.001** |
| Undetermined | 172 (2.43) | 194 (2.29) | 0.571 |
| **Hospitalization** |  |  |  |
| Hospitalization days, median (IQR) | 8 (5-15) | 8 (5-15) | 0.945 |
| Stroke care unit<sup>c</sup> | 2,822 (39.89) | 3,347 (39.56) | 0.671 |
| Mechanical ventilation | 686 (9.70) | 747 (8.83) | 0.062 |
| Readmission | 224 (3.17) | 297 (3.51) | 0.236 |
| **Diagnostic test** |  |  |  |
| Brain CT | 6,513 (92.07) | 7,820 (92.42) | 0.410 |
| Brain MRI | 824 (11.65) | 1,229 (14.53) | **<0.001** |
| In-hospital complications<sup>d</sup> | 1,873 (26.48) | 2,201 (26.01) | 0.513 |
| Dysphagia | 634 (8.96) | 699 (8.26) | 0.120 |
| Pneumonia | 825 (11.66) | 1,151 (13.60) | **<0.001** |
| Pressure ulcers | 288 (4.07) | 271 (3.20) | **0.004** |
| ARDS | 164 (2.32) | 215 (2.54) | 0.370 |
| Urinary tract infection | 34 (0.48) | 23 (0.27) | **0.032** |
| Acute kidney injury | 286 (4.04) | 436 (5.15) | **0.001** |
| In-hospital mortality | 1,172 (16.57) | 1,266 (14.96) | **0.006** |

Abbreviations: IQR=interquartile range; SD= standard deviation; OACs= direct oral anticoagulants; MRI= magnetic resonance imaging; CT= computed tomography; ARDS= acute respiratory distress syndrome.<sup>a</sup>Among those with National Healthcare insurance <sup>b</sup>Two or more comorbidities. <sup>c</sup>Measured at hospital level. <sup>d</sup>At least one in-hospital complication. Bold denotes statistically significant results.

Women accounted for 45.54% (7,074/15,535) of stroke and they were older than men (70.4±14.8 years versus 67.0±13. 5 years; *P*<0.001). Table 1 outlines the other patient characteristics, stroke care and outcomes. After adjustment for age, women had lower SES (OR 1.92, 95%CI1.77-2.08; *P*<0.001), had more HT (1.10, 1.01-1.18; *P*=0.01), obesity (1.64, 1.45-1.86; *P*<0.001), and valvular heart diseases (1.28, 1.05-1.55; *P*=0.01) than men (Table S2). Conversely, women were less often smokers (0.64, 0.57-0.71; *P*<0.001) and with a history of ischemic heart disease (IHD) (0.53, 0.46-0.60; *P*<0.001) and they received less use of magnetic resonance imaging (MRI) (0.82 0.75-0.91; *P*<0.001). In-hospital complications by sex showed lower pneumonia (0.75, 0.68-0.83; *P*<0.001) and acute kidney injury (0.73, 0.62-0.85; *P*<0.001) for women than men (Table S2). Overall, 2,438 (15.69%) patients died during hospitalization.

### Ischemic Stroke

Among 10,440 patients with IS, 5,781 were men (55.37%) who tended to be younger than women (67.9±12.6 years versus 71.6±14.2 years; *P<*0.001); Table 2 defines other characteristics and outcomes. After adjustment for age, women had lower SES (2.01, 1.81-2.22; *P*<0.001) and more risk factors of HT (1.16, 1.05-1.28; *P*=0.002), diabetes mellitus (DM) (1.09, 1.01-1.18; *P*=0.02), obesity (1.74, 1.49-2.02; *P*<0.001), valvular heart diseases (1.39, 1.12-1.71; *P*=0.002) and oral anticoagulant (OAC) use (1.18, 1.001-1.38; *P*=0.03). However, women had less IHD (0.56, 0.48-0.65; *P*<0.001) and current smoking (0.55, 0.49-0.63; *P*<0.001) than men (Figure 2). There were no significant sex differences in the duration of hospitalization (*P*=0.12), the availability of a stroke unit (*P*=0.57) or the use of reperfusion treatment, whether intravenous thrombolysis (*P*=0.46), thrombectomy (*P*=0.91) or craniectomy (*P*=0.32). Regarding imaging, although there was no sex-difference in the use of computerized tomography (CT) (*P*=0.94), women had less MRI (0.80, 0.72-0.89; *P*<0.001) than men. Regarding in-hospital complications, women had less pneumonia (0.66, 0.58-0.75; *P*<0.001) but more urinary tract infections (UTI) (2.35, 1.16-4.75; *P*=0.01) compared to men (Figure 2).

**Figure 2.**
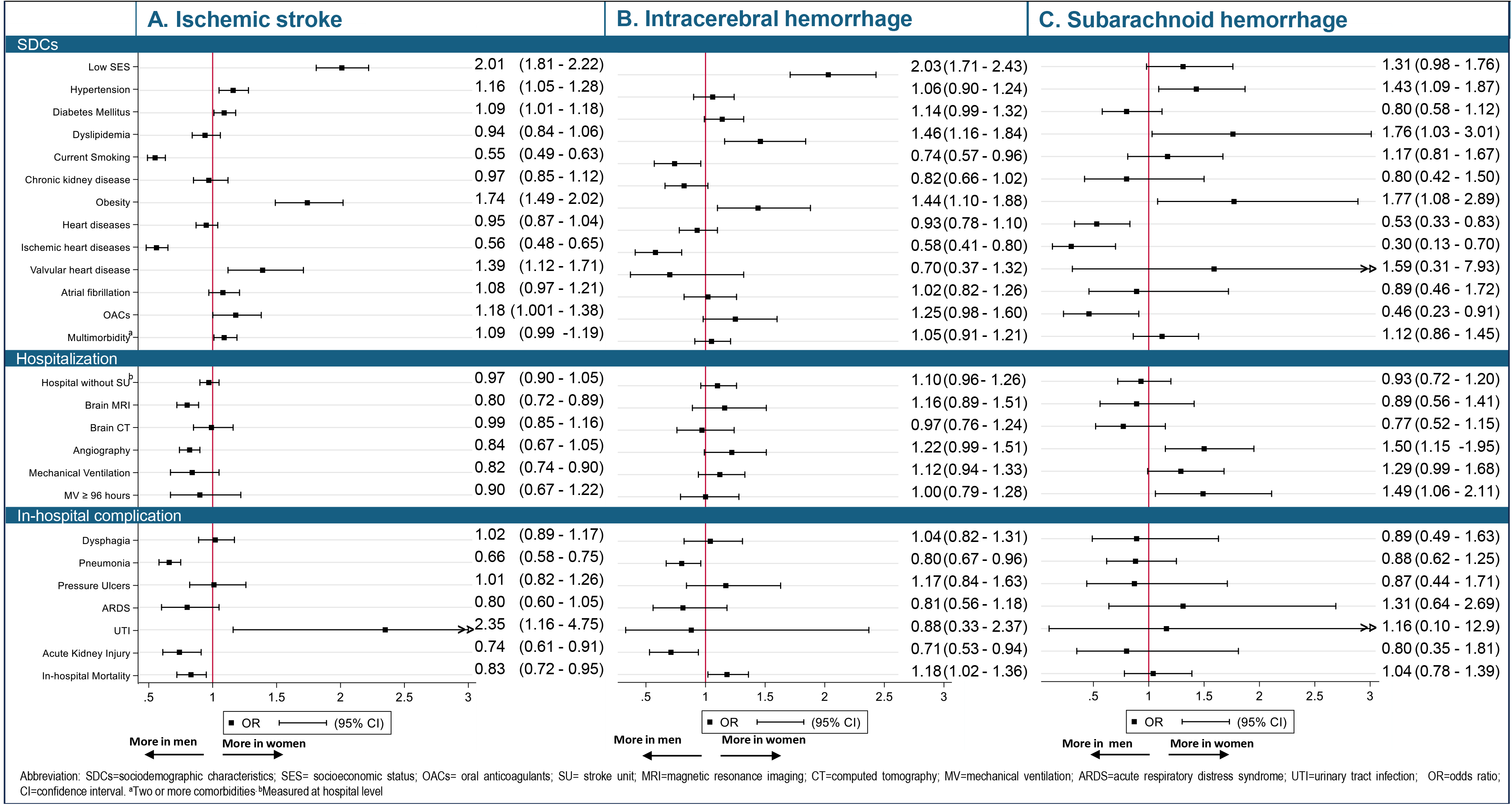
Age-adjusted odds ratio of baseline characteristics by stroke subtype in women compared to men.

**Table 2.**
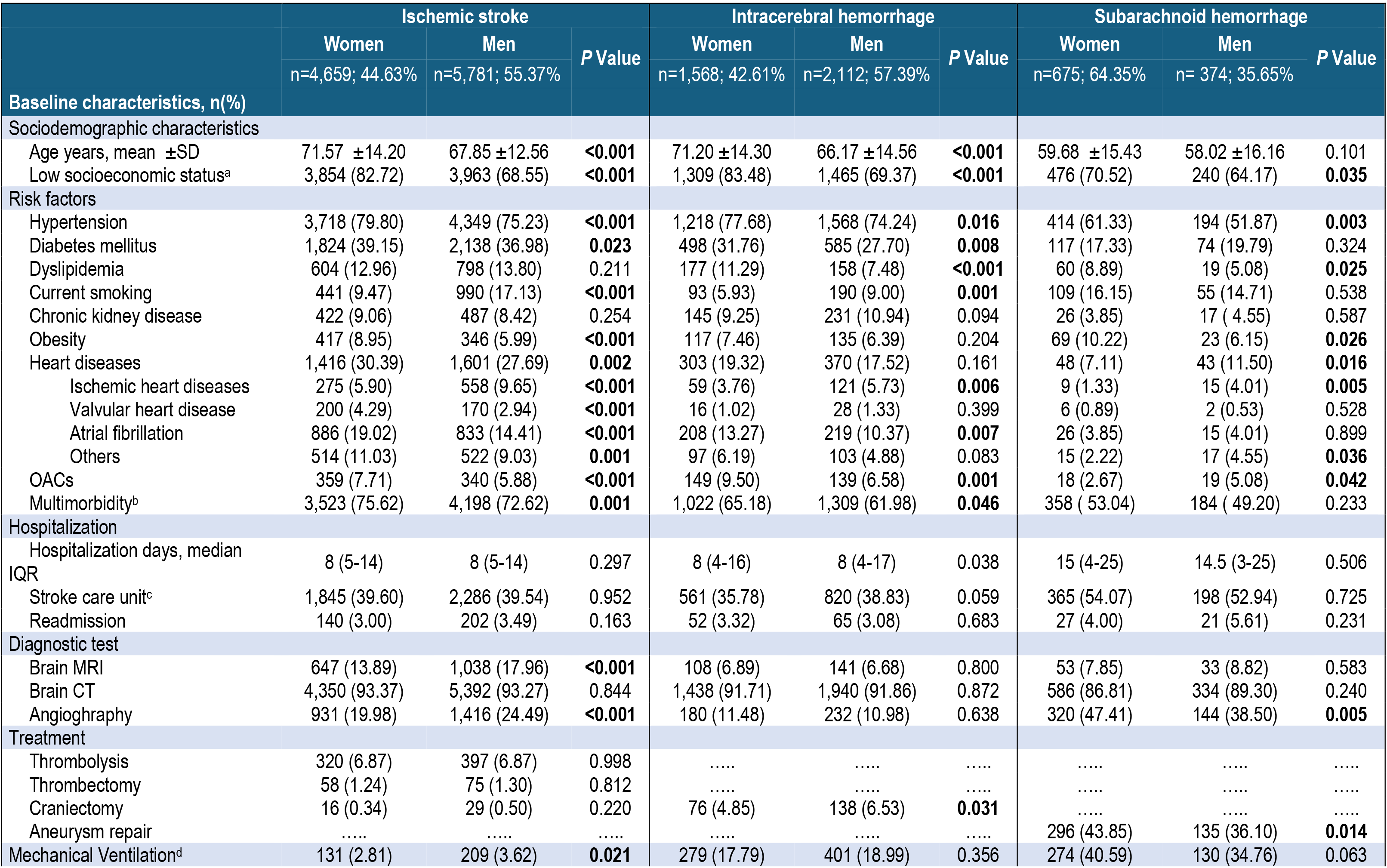
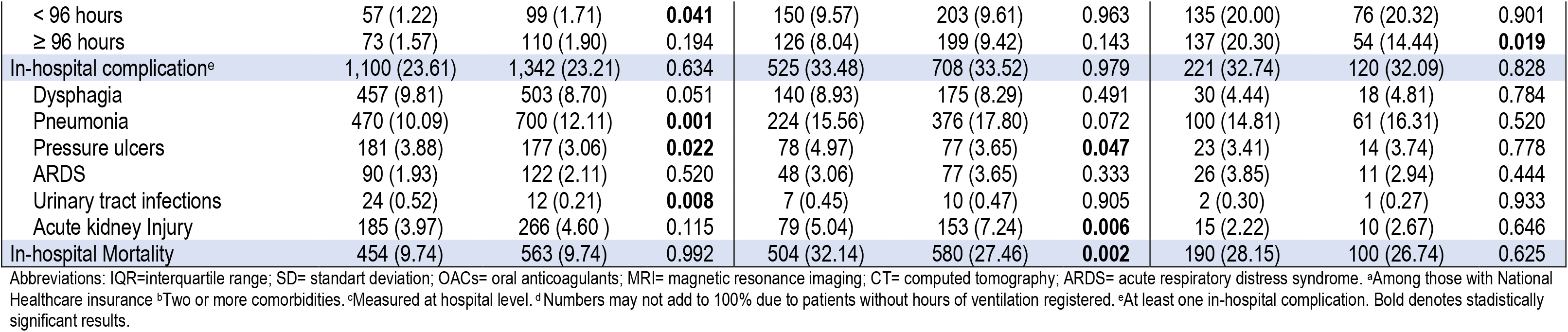
Characteristics and outcome of acute stroke patients according to stroke subtype by sex.

### In-Hospital Mortality in Ischemic Stroke (I-HM_IS_)

There were 1,017 (9.74%) patients who died in hospital but there was no significant differences in the crude I-HM_IS_ between the sexes. However, after adjustment for age, women had a lower I-HM_IS_ than men (0.83, 0.72-0.95; *P*=0.008) (Figure 2) and this result remained significant in the fully adjusted model (0.80, 0.70-0.92; *P*=0.002) (Tables S3 and S4, Figure 3). Other variables independently associated with I-HM_IS_ were age (1.03, 1.03-1.04; *P*<0.001), CKD (1.48, 1.20-1.81; *P*<0.001), AF (1.56, 1.34-1.83; *P*<0.001), admission to a hospital without SU (1.21, 1.05-1.39; *P*=0.003), dyslipidemia (0.40, 0.31-0.53; *P*<0.001), current smoking (0.66, 0.51-0.86; *P*=0.002) and the presence of valvular heart disease (0.52, 0.33-0.80; *P*=0.003). Interestingly, being admitted to a hospital without stroke unit was especially relevant for men with IS (1.27, 1.05-1.54; *P*=0.013) but not women (Table S4). Other sex disaggregated predictors of I-HM_IS_ are outlined in Supplementary Table S4.

**Figure 3.**
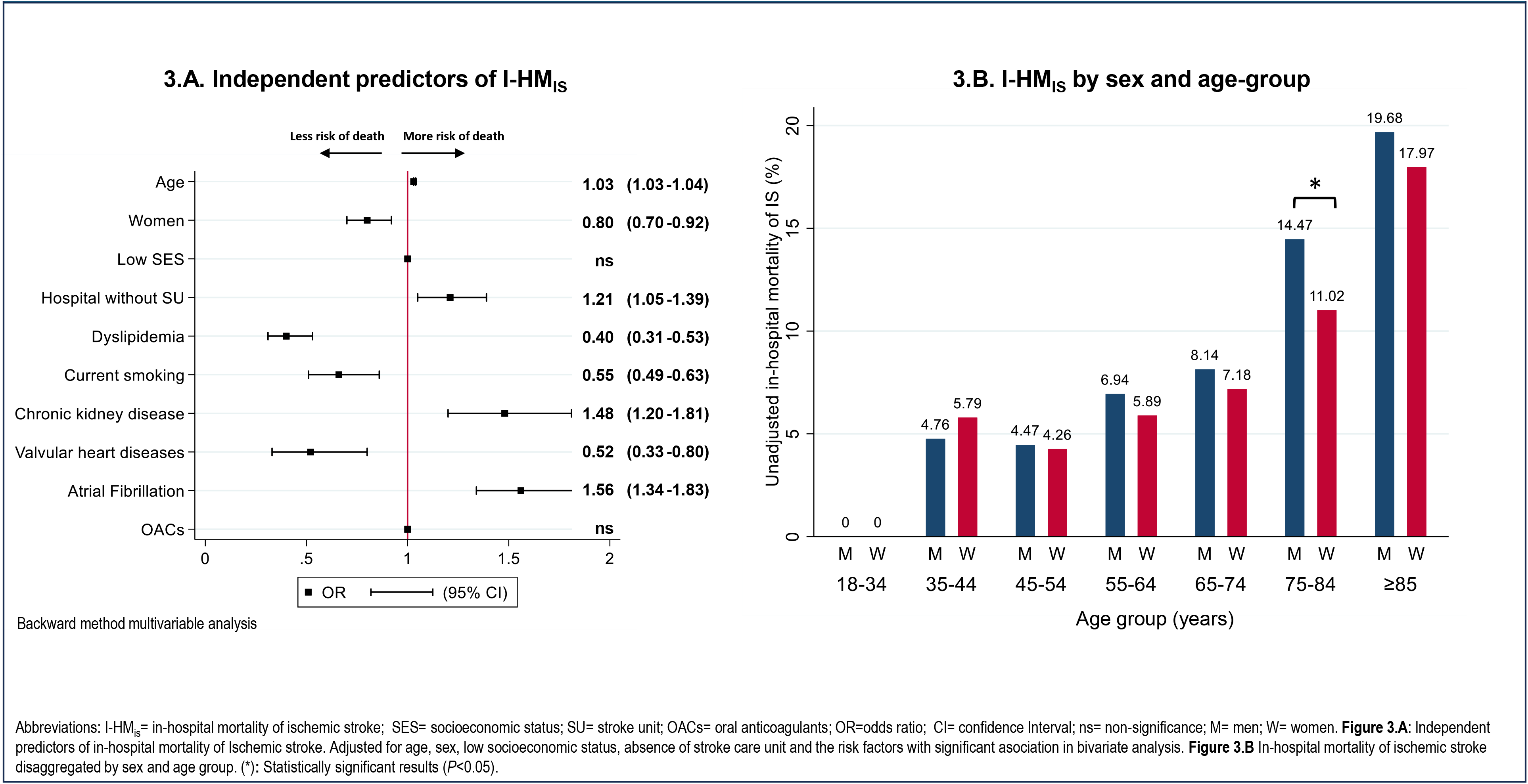
Predictors of in-hospital mortality of ischemic stroke, by sex and age-group.

Women had lower risk of dying between only between the ages of 75 and 84 years in the fully adjusted analysis when compared to men (0.70 [CI, 0.56-0.89]; *P*=0.004) (Figure 3, Table S5).

### Intracerebral hemorrhage (ICH)

Of 3,680 ICH patients, 2,112 were men (57.39%) who were younger than women (66.2±14.6 years versus 71.2±14.3 years)*; P<*0.001) (Table 2). Adjustment for age showed that women had lower SES (2.03, 1.71-2.43; *P*<0.001) and more dyslipidemia (1.46, 1.16-1.84; *P*<0.001) but less current smoking (0.74, 0.57-0.96; *P*=0.02) and history of IHD (0.58, 0.41-0.80; *P*=0.001). Women also had less acute kidney injury (AKI) compared to men (0.71, 0.53-0.94; *P*=0.01) (Figure 2). There were no significant sex differences in the duration of hospitalization (*P*=0.08), the availability of a stroke unit (*P*=0.15), the use of CT scan (*P*=0.84), and craniectomy (P=0.08).

### In-hospital mortality in ICH (I-HM_ICH_)

There were 1,084 (29.46%) ICH patients who had an in-hospital death. Women had higher I-HM_ICH_ than men (32.1% versus 27.5%; age-adjusted OR 1.18, 95%CI 1.02-1.36 [*P*=0.02)] and fully-adjusted (OR 1.21, 1.04-1.41 [*P*=0.01]) (Table S3 and Table S6). Other independent predictors of I-HM_ICH_ were age (1.01, 1.00-1.01; *P*<0.001), CKD (1.54, 1.22-1.94; *P*<0.001), OAC use (2.01, 1.56-2.59; *P*<0.001), HT (0.72, 0.60-0.85; *P*<0.001), current smoking (0.63, 0.45-0.86; *P*=0.004) and dyslipidemia (0.49, 0.36-0.66; *P*<0.001). Unlike women, chronic kidney disease was associated to I-HM_ICH_ in men (1.99, 1.48-2.68, P<0.001). This and other sex-disaggregated predictors of I-HM_ICH_ are outlined in Tables S6 and Figure 4.

**Figure 4.**
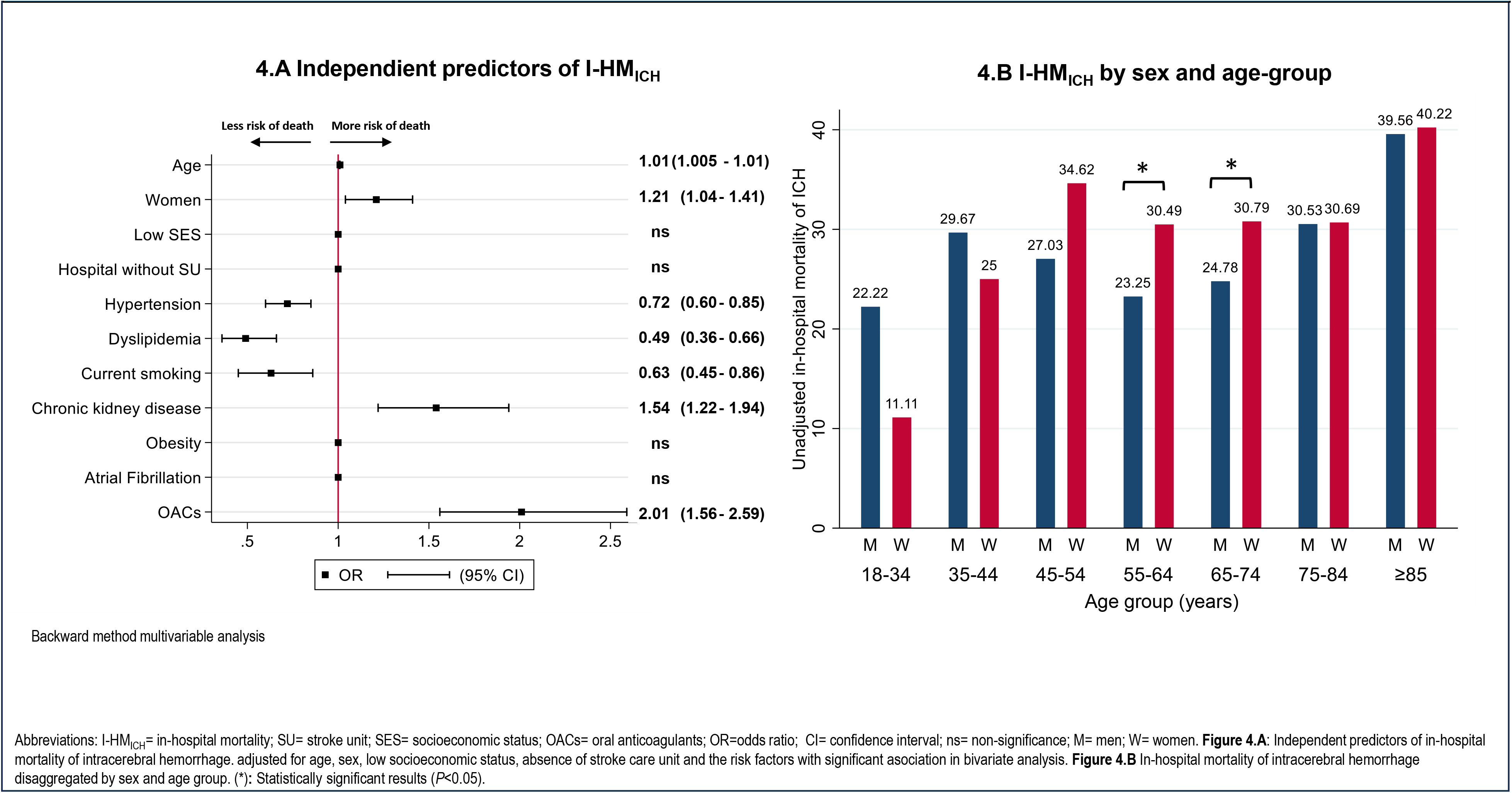
Predictors of in-hospital mortality for intracerebral hemorrhage, by sex and age-group.

Women with ICH between the ages of 55-64 and 65-74 years had significantly higher I-HM_ICH_ than men (Figure 4,Table S5). In the fully adjusted model, the age range 55-74 years remains significant (1.42, 1.12-1.79; *P*=0.003).

### Subarachnoid hemorrhage (SAH)

Of 1,049 patients hospitalized for SAH, 675 were women (64.35%); there was no age difference by sex (women 59.7±15.4 years versus men 58.0±16.2 years; *P=*0.10) (Table 2). After adjustment for age, women had more HT (1.43, 1.09-1.87; *P*=0.01), dyslipidemia (1.76,1.03-3.01; *P*=0.03) and obesity (1.77, 1.08-2.89; *P*=0.02), but less IHD (0.30, 0.13-0.70; *P*=0.005) and OAC use (0.46, 0.23-0.91; *P*=0.02), than men. Women had more angiography (1.50, 1.15-1.95; *P*=0.002) and more aneurysm repair procedures than men (1.47, 1.12-1.92; *P*=0.004) (Figure 2). There were no significant sex differences in the duration of hospitalization (*P*=0.77), the availability of a stroke unit (*P*=0.60) and in the use of CT scan (*P*=0.21).

### In-hospital mortality in SAH (I-HM_SAH_)

There were 290 (27.65%) patients who died of a SAH. For I-HM_SAH_, there was no difference by sex (1.07, 0.80-1.42) even with adjustment for age (1.04, 0.78-1.39) (Figure 2). Independent predictors of I-HM_SAH_ were age (1.01, 1.007-1.02; *P*<0.001) and admission to a hospital without a stroke unit (1.37, 1.04-1.82; *P*=0.025). In contrast to the factors for IS, lack of an in-hospital stroke unit was associated with I-HM_SAH_ for women but not men (1.56, 1.10-2.20; *P*=0.011) (Table S7). Other sex disaggregated predictors of I-HM_SAH_ can be found in Supplementary Table S7.

## Discussion

In this study, which used national data for annual hospital discharges for stroke, we outline sex disaggregated data for sociodemographic, clinical characteristics and outcome. We show that sex influences I-HM by stroke subtype, that is higher in men with IS and women with ICH.

The characteristics of our patients with acute stroke is generally consistent with other populations, in that women tend to be older, of lower SES, and have greater comorbidities of HT, AF, and obesity but less ischemic heart disease and smoking, than men.^2,6,16–19^ Although we were unable to account for baseline severity in our analysis, our observations align with other studies in showing a greater survival in women after IS.^5,16,20–24^ The reasons for this remain unexplained, but raise issues of sexual dimorphism in neuroanatomy or biology,^21^ unequal interaction of predictor variables,^23^ and a survival advantage in relation to post-stroke infections.^24^ A Japanese Stroke Registry of 19,956 IS patients showed that women had a lower-case fatality among those who developed a post-stroke infection.^24^ Our finding of impact of stroke unit care on short term mortality in men with IS could not be adequately explained on the basis of differential stroke severity. Further research is needed to understand underlying factors sex disparities on survival after IS.

For ICH, women were 21% more likely to die in hospital than men, and this could not be explained by age or baseline risk factors. Significantly, this distinction was noted during a productive phase of life, especially among individuals aged 55 and older. While conflicting results shown elsewhere^6–8,17,25^ may stem from differences in methodologies, there may also be sociocultural explanations.^25,26^ Women in our study where twice as likely to belong to a lower SES than men, and although this was controlled in the multivariable analysis, it is important to note that disadvantaged women are more susceptible to gender-based violence, social isolation, poor social support, and insufficient education than men.^27–30^ A the 479,054 participants of the UK biobank, social isolation but not loneliness was associated with higher risk of all-cause mortality.^31^ A national health survey 4,473 people in Chile showed poorer health-related quality of life reported in women than men.^32^ Thus, the social context of Chilean women may contribute to a higher probability of a severe presentation and worse outcome in stroke.

Furthermore, early mortality is inherently connected with the quality of hospital care,^33–35^ but the influence of sex-based bias in care is under-reported. In cardiovascular studies, men are often perceived as being physically stronger, leading healthcare providers to be more inclined to recommend and perform medical procedures for them than women.^36^ There are few reports specifically in relation to the management of patients with acute ICH where a performance focus is on do not resuscitate (DNR) orders: results suggest higher use of DNR in women than men,^37^ although such differences could relate to incomplete adjustment for confounding variables.^38^ ICH patients with severe illness are unable to make critical end-of-life decisions. While we did not have access to the severity of the initial illness or data on various quality metrics, given the strong connection between in-hospital mortality and quality of care, it is plausible that the greater severity of ICH in women and the perception of them being more fragile might have influenced decisions. Unconscious gender-related biases may also have influenced the findings of fewer medical interventions in women, which potentially limited diagnostic and therapeutic efforts. The revealed trend of lower number of craniectomies in women compared to men, that was not significant after age adjustment, might be interesting to explore in a cohort with more surgeries in ICH. Further investigation into sex disparities in the intensity of care after acute ICH is clearly warranted.

Our study has several limitations. In particular, we lacked data on the time from the onset of symptoms to presentation, severity of the neurological deficit and the place of care, and other management that are associated with outcomes. There might have been miscoding errors and missing data that influenced the results. However, the large sample size, national representation,^13^ and non-differential bias in the data, still provides a reliable overview of sex disparities in the early outcome from acute stroke in Chile.

In summary, we have defined sex differences in the characteristics, management and early I-HM for patients with acute stroke in Chile. The impact of sex on I-HM across the subtypes of stroke may stem from differences in the social determinants of health and unconscious bias in the delivery of care. Further research is required to provide better granularity over factors contributing to sex differences in early stroke mortality.

## Data Availability

Data that support the findings of this study are available from the corresponding author upon reasonable request.

## Acknowledgment

None.

